# Prevalence, trends, and determinants of malnutrition among under six-month-old infants in Bangladesh: analysis of DHS data (2004 – 2022)

**DOI:** 10.64898/2026.02.10.26345991

**Authors:** Imteaz Mahmud, Roxanne Assies, Rukhsana Haider, Azaz Bin Sharif, Kedir Teji Roba, Marko Kerac

## Abstract

Malnutrition among infants aged under six months (u6m) is a global public health problem. As countries begin implementing 2023 WHO malnutrition guidelines, local prevalence and epidemiology must be well understood. We analysed Bangladesh Demographic Health Survey data (2004 – 2022), describing the prevalence and time trends of infant u6m wasting (weight-for-length z-score [WLZ] <-2), stunting (length-for-age z-score [LAZ] <-2), and underweight (weight-for-age z-score [WAZ] <-2). In bivariate and multivariate analyses, we tested the association between wasting and underweight and established risk factors for malnutrition. Over the last 20 years, Bangladesh has seen a substantial reduction in stunting and underweight while wasting has remained relatively stable. In 2022, out of 476 infants u6m, 10.6% were wasted, 12.8% stunted, 13.7% underweight, 0.5% concurrently wasted/stunted, and 14.8% were reported small at birth. This translates to 185,390 infants u6m being wasted, 223,867 stunted, and 239,608 underweight, in 2022. WAZ had the lowest percentage of flagged data (0.8%) compared to WLZ (6.0%) and LAZ (6.4%). Underweight was associated with delivery place, small birth size, infant sex, post-natal check-ups, fever in the past 2 weeks, drinking water source, maternal BMI, and maternal height. Fewer factors were associated with being wasted, including the sex of the household head. Our findings support the urgent need to roll out 2023 WHO Guidelines in Bangladesh. They also support the superiority of underweight as a measure of undernutrition. Packages of care tackling a wide range of potential underlying causes are important for effective local interventions tailored to this age group.

## 1 INTRODUCTION

Malnutrition remains a major global public health challenge, disproportionately affecting low- and middle-income countries (LMICs), particularly in Africa and Asia (Marko et al., 2020; Organization, 2023). Approximately 2.4 million under-five deaths are attributed to malnutrition annually, which is half of all global child deaths (Ritchie, 2024). Among survivors, malnutrition has long-term consequences, including impaired physical and cognitive development (Amir et al., 2022; Bhutta et al., 2017; Black et al., 2013) and an increased risk of later-life non-communicable diseases (Kelsey et al., 2021; Lelijveld et al., 2016). According to the 2025 Joint Child Malnutrition Estimates, an estimated 150.2 million (23.2%) children under five years of age are stunted, and 42.8 million (6.6%) are wasted globally (United Nations Children’s Fund (UNICEF), 2025).

Infants aged under six months (u6m) are increasingly recognised as a highly vulnerable subgroup. A recent analysis of 56 Demographic and Health Surveys (DHS) estimated that 10.3 million (17.4%) infants u6m are underweight, 9.2 million (15.5%) are wasted, 11.8 million (19.9%) are stunted, and 8.9 million (15.0%) are born with low birth weight (Kerac et al., 2025).

Sustainable Development Goal (SDG) 2, Zero Hunger, underscores the critical importance of addressing malnutrition (*Sustainable Development Goals: Goal 2: Zero Hunger*). However, progress toward achieving a 50% reduction in undernutrition by 2030 has been severely disrupted by the COVID-19 pandemic, global conflicts, and climate crises (Nations, 2022). To counteract this setback, a more aggressive, effective and targeted approach is needed, particularly focusing on infants u6m, who have been historically overlooked in nutrition policy and programming. The World Health Organisation (WHO) recently emphasised the importance of infants u6m by including them as a key group in the new 2023 wasting guideline (WHO, 2024). Termed ‘infants at risk of poor growth and development’, the criteria for programme enrolment now go beyond wasting alone and include low weight-for-age z-score (WAZ < -2), low weight-for-length z-score (WLZ < -2), and low mid-upper arm circumference (MUAC <110 mm, for infants aged over 6 weeks to 6 months) (WHO, 2024). Whilst a major step forward, that infants u6m are emphasised in the new guideline, the recommendations are based on conditional and low-certainty evidence. Robust data from diverse settings is needed to strengthen policies and intervention strategies. Among the key questions arising is which anthropometric and other criteria should be used to identify the highest-risk infants u6m for priority enrolment and treatment. There has been particular recent interest in possible advantages of weight-for-age over weight-for-length as an anthropometric measure best able to identify the highest-risk infants (Ahmed et al., 2025; Hoehn et al., 2021).

Bangladesh, one of the most densely populated countries in the world, faces a significant burden of malnutrition, with 24% of children aged under five years stunted, 22% underweight, and 11% wasted (ICF, 2023). Despite ongoing public health efforts, child malnutrition persists due to a complex interplay of poverty, food insecurity, inadequate water, sanitation, and hygiene (WASH) practices, frequent natural disasters, suboptimal maternal and child health practices, and limited healthcare access (Faruque et al., 2008; Mondal et al., 2024; Nabeela Ahmed). While much research has focused on malnutrition in older children, data specific to infants u6m remain scarce.

In this study, we use the nationally representative dataset of the last two decades from Bangladesh to:

- Assess the prevalence and concurrence of different forms of anthropometric deficit in infants u6m
- Evaluate the quality of anthropometric data
- Understand the determinants of infant u6m malnutrition in Bangladesh

As many countries worldwide seek to implement the 2023 WHO Malnutrition guideline, our approach to understanding local prevalence and epidemiology of infant u6m malnutrition also aims to provide a useful model for others to follow.

### Key messages

- In Bangladesh, the prevalence of wasting, stunting, and underweight among infants u6m has reduced over the last 20 years, though it is still high.
- WAZ shows better data quality and stronger coherence with epidemiology than WLZ and LAZ. It supports greater practicality for routine surveys and monitoring nutrition programmes.
- A strong association between infant underweight and maternal anthropometry, access to care, and size at birth highlights the importance of maternal and perinatal nutrition in preventing infant malnutrition.
- This high burden of co-existing malnutrition among infants u6m underscores the need for an integrated mother-newborn-infant nutrition strategy.

## 2 METHODS

### 2.1 Study design

In this secondary analysis, we used the Bangladesh Demographic and Health Surveys (BDHS) dataset from 2004 to 2022 (ICF, 2025). The DHS program has conducted standardised household surveys in multiple countries since 1984, providing reliable data on health and nutrition. In Bangladesh, these cross-sectional, nationally representative surveys were conducted every three to four years from 1993 to 2022. This dataset is publicly available. We got access to this dataset on 28^th^ January, 2022. As part of the original survey protocol, all data were anonymised prior to download from repositories to protect participant privacy. This study focuses on infants u6m, a population group that has received limited attention in nutrition research despite its recognised vulnerability.

### 2.2 Sampling strategy and data collection

The BDHS employed a two-stage cluster sampling strategy to ensure a nationally representative sample (NIPORT, 2024). The sampling frame was based on the most recent census Enumeration Areas (EAs), each containing approximately 120 households serving as Primary Sampling Units (PSUs). In stage one, EAs were stratified by urban and rural residences, and a random selection of PSUs was made using probability proportional to size. An average of 30 households per EA were randomly selected for data collection in stage two.

Trained interviewers visited selected households and collected demographic and anthropometric data using the DHS standard household questionnaire. Children under 5 years were identified, and their weight and length were measured using standardised procedures. Weight was measured using a digital scale to the nearest 0.1 kg, and length was measured using a length board, with children <2 years or <85 cm measured lying down. Additional maternal and household information was collected during the survey.

### 2.3 Variables

Infants u6m were identified from the children’s dataset. Weight (kg) and length (cm) were measured to one decimal place. Data quality checks included generating histograms and scatter plots to assess distribution and identify outliers. Anthropometric indices (z-scores) were calculated using the WHO growth reference standards via the zscore06 package (Leroy, 2011). The following cut-offs were applied for biologically implausible values (Unicef, 2009):

- Weight-for-length z-score (WLZ): < -5 or > 5
- Length-for-age z-score (LAZ): < -6 or > 6
- Weight-for-age z-score (WAZ): < -6 or > 5

Nutritional status was classified as follows:

- Wasting: WLZ < -2
- Stunting: LAZ < -2
- Underweight: WAZ < -2
- Severe wasting/stunting/underweight: WLZ/LAZ/WAZ < -3
- Moderate wasting/stunting/underweight: WLZ/LAZ/WAZ between -2 and -3
- Concurrent wasting and stunting (WaSt): WLZ < -2 & LAZ < -2

To examine determinants of malnutrition, we selected key infant, maternal, and household factors based on UNICEF’s Analytical Framework for COVID-19 and Malnutrition (*The Analytical Framework for COVID-19 and nutrition*). Variables were recoded according to established definitions. For instance, the preceding birth interval was categorised as <36 months vs. ≥36 months, based on prior literature (Rutstein, 2005), and exclusive breastfeeding was defined according to the WHO criteria (Unicef, 2021). Explanatory variables were classified into infant, maternal, and household characteristics. Infant characteristics included age (<1month / 1 to <2 months /2 to <4 months/4 to <6 months), sex (male/female), birth weight (reported size at birth) (average or large/small), birth spacing (<=36 months/> 36 months), number of antenatal care (ANC) visits (<4/>=4), place of delivery (home/health facility), received postnatal care (PNC) within 2 months of delivery (yes/no), time of breastfeeding initiation (< 1 hour/> 1 hour), current breastfeeding status (yes/no), exclusive breastfeeding (yes/no), bottle feeding (yes/no), infants given powdered, tinned or fresh milk (Yes/No), recent illness (cough, fever, diarrhoea) (yes/no) and presence of vaccine card (yes/no). Maternal characteristics included maternal age at delivery (<20 year/20-24 years/>=35 years), education level (no education/primary/secondary/higher), maternal height (<145cm/>=145cm), BMI (normal/underweight/overweight or obese), employment status (employed/unemployed), and total births (< 3 / >=3). Household characteristics included residence type (urban or rural), sex of the household head (male/female), religion (Muslim/Hindu and others), drinking water source (improved/unimproved), cooking fuel (clean/unclean), toilet facilities (improved/unimproved), wealth quintiles (poorest/poorer/middle/richer/richest). All variables were captured using standard DHS methodologies, with detailed descriptions available in the official DHS manual (*DHS Manuals*).

### 2.4 Statistical analysis

We performed statistical analysis using STATA 16 (StataCorp, 2017). Sampling weights were used to estimate frequencies and percentages to adjust for under- and oversampling. To account for the complex survey design, we applied svyset command to obtain robust standard errors. We explored the distribution of different anthropometric measurements of malnutrition and their concurrence across age groups using the combined 2004 to 2022 dataset. We assessed the prevalence of malnutrition indicators (wasting, stunting, underweight, and WaSt) across 2004, 2007, 2011, 2014, 2018, and 2022 to evaluate trends over time. Based on the latest 2022 prevalence, we estimated the current caseload of malnutrition within this age group in Bangladesh. Using combined data (2004 – 2022), we examined crude associations (unadjusted odds ratios) between infant nutritional status (wasted and underweight) and associated factors using bivariate logistic regression. Finally, we performed multivariable logistic regression to find the determinants of malnutrition among infants u6m. All the numbers mentioned here were based on weighted analysis, except the study participants flow chart.

### 2.5 Ethical considerations

This study used publicly available, de-identified data from the BDHS, accessed with permission from the DHS Program. The BDHS surveys are ethically approved by the Institutional Review Board (IRB) of ICF International and the Bangladesh Medical Research Council (BMRC). The project was further approved by the LSHTM Research Ethics Committee (reference 15051).

## 3 RESULTS

### 3.1 Study participants

**Figure 1** shows the participant’s flow chart. 6 BDHS rounds datasets (2004 – 2022) were included in this analysis. After excluding infants with missing or flagged data, a total of 3,625 infants u6m were included in this study.

**Figure 1:**
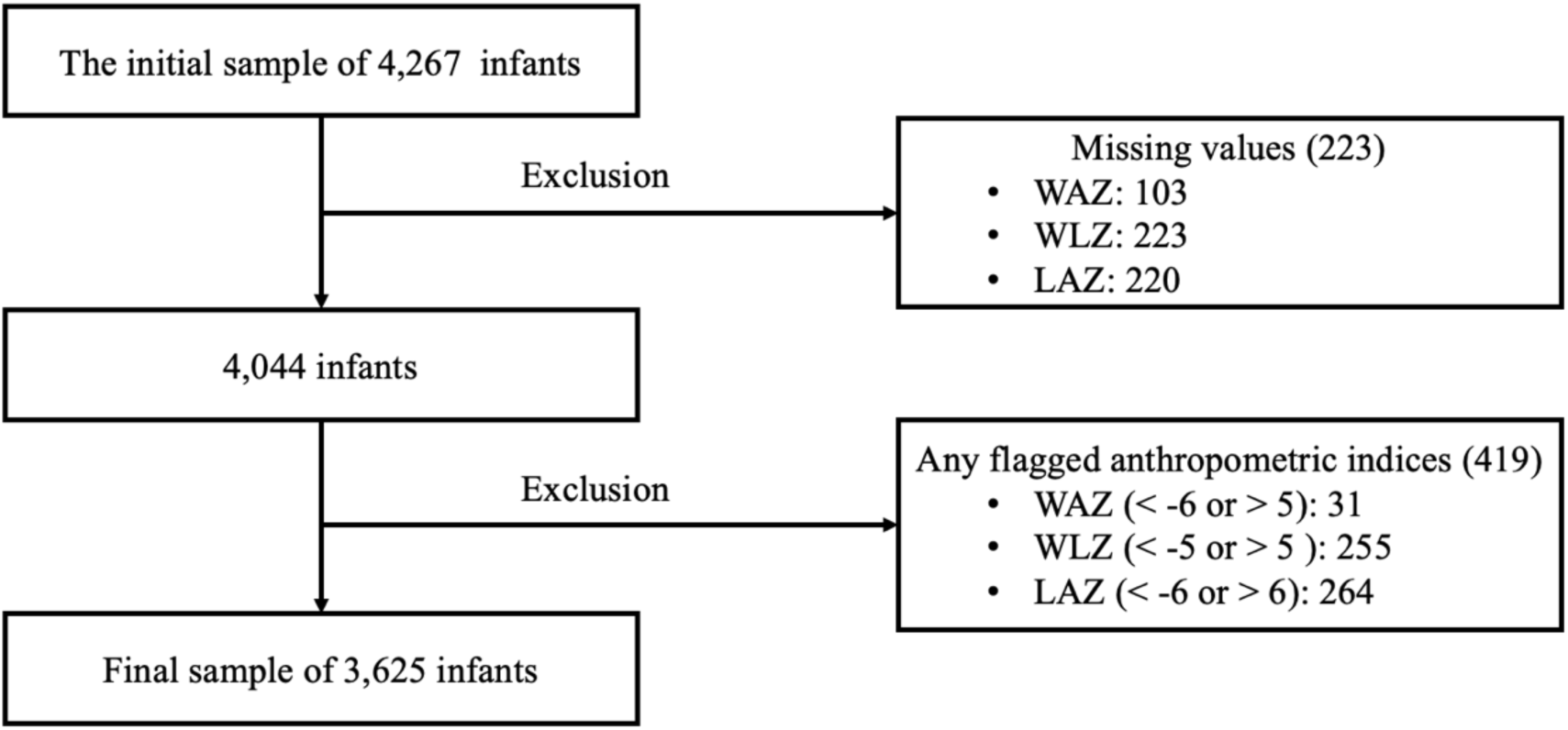
Study participants’ flow chart (not weighted). WAZ, weight-for-age *z*-score; WLZ, weight-for-length/height *z*-score; LAZ, length-for-age *z*-score.

The distribution of the sample population across the eight districts of Bangladesh is shown in **Table S1**. Around half of the participants were from the Dhaka and Chattogram divisions. Sylhet division had the lowest representation (5.4%) in this analysis.

### 3.2 Quality of the data

**Table 1** shows the distribution of flagged (out-of-range) data among the collected sample. LAZ had more flagged values (n=259, 6.4%) than WLZ (n=243, 6.0%) and WAZ (n=34, 0.8%). A total of 401(10.0%) participants had any form of flagged data.

**Table 1:**
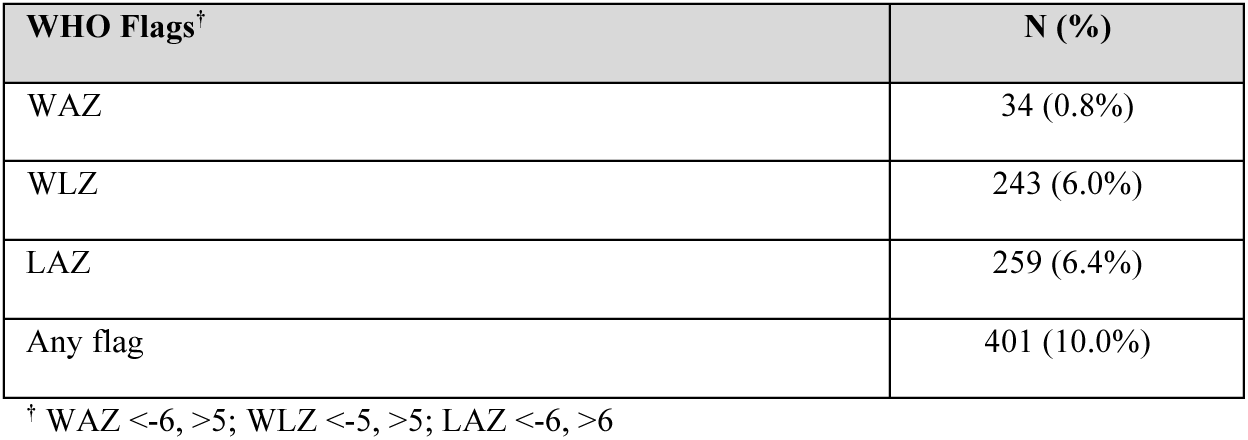
Distribution of flagged data.

| WHO Flags<sup>†</sup> | N (%) |
| --- | --- |
| WAZ | 34 (0.8%) |
| WLZ | 243 (6.0%) |
| LAZ | 259 (6.4%) |
| Any flag | 401 (10.0%) |

<sup>†</sup> WAZ <-6, >5; WLZ <-5, >5; LAZ <-6, >6

### 3.3 Distribution of different anthropometric deficits

**Table 2** shows the distribution of infants by age in months, sex, and different anthropometric deficits, including moderate and severe wasting, stunting, underweight, concurrence of wasting and stunting and reported small at birth.

**Table 2:** Descriptive data for anthropometric measurements by infant age category.

| Characteristics |  | Age of the infants (months) (Year 2004 – 2022) |  |  |  |  | Year 2022 | Total national caseload in Bangladesh (N)<sup>‡</sup> |
| --- | --- | --- | --- | --- | --- | --- | --- | --- |
|  |  | <1m | 1 m | 2 to <4 m | 4 to <6 m | Total | Total |  |
| Number of Infants | N (%) | 550 (15.1) | 560 (15.4) | 1,208 (33.3) | 1,311 (36.1) | 3,629 (100) | 476 (100) | 1,748,963 |
| Sex (female) | N (%) | 273 (49.6) | 273 (48.8) | 587 (48.6) | 635 (48.5) | 1,768 (48.7) | 233 (48.9) | 855,243 |
| Weight (kg) | Mean (SD) | 3.4 (0.6) | 4.3 (0.7) | 5.4 (0.8) | 6.5 (0.9) | 5.3 (1.4) | 5.22 (1.5) |  |
| Length (cm) | Mean (SD) | 50.7 (2.5) | 54.7 (2.7) | 59.1 (2.8) | 63.3 (2.9) | 58.6 (5.2) | 58.2 (5.7) |  |
| Underweight |  |  |  |  |  |  |  |  |
| Mean WAZ | Mean (SD) | -0.9 (1.0) | -1.0 (1.1) | -0.9 (1.1) | -0.9 (1.1) | -0.9 (1.1) | -0.9 (1.1) |  |
| Underweight | N (%) | 66 (12.0) | 102 (18.2) | 191 (15.8) | 217 (16.5) | 576 (15.9) | 64 (13.7) | 239,608 |
| Moderate underweight | N (%) | 59 (10.8) | 81 (14.4) | 158 (13.1) | 175 (13.3) | 473 (13.0) | 46 (9.7) | 166,999 |
| Severe underweight | N (%) | 7 (1.1) | 21 (3.8) | 33 (2.7) | 42 (3.2) | 103 (2.8) | 19 (4.0) | 69,959 |
| Wasting |  |  |  |  |  |  |  |  |
| Mean WLZ | Mean (SD) | -0.6 (1.3) | -0.6 (1.3) | -0.5 (1.2) | -0.5 (1.2) | -0.5 (1.2) | -0.4 (1.3) |  |
| Wasted | N (%) | 85 (15.4) | 78 (14.0) | 157 (13.0) | 139 (10.6) | 459 (12.6) | 47 (10.6) | 185,390 |
| Moderate wasted | N (%) | 68 (12.4) | 60 (10.8) | 138 (11.4) | 116 (8.9) | 382 (10.5) | 42 (9.4) | 164,577 |
| Severe wasted | N (%) | 17 (3.0) | 18 (3.2) | 19 (1.6) | 23 (1.7) | 77 (2.1) | 5 (1.2) | 20,813 |
| Stunting |  |  |  |  |  |  |  |  |
| Mean LAZ | Mean (SD) | -0.8 (1.1) | -0.6 (1.2) | -0.6 (1.1) | -0.7 (1.2) | -0.7 (1.2) | -0.6 (1.3) |  |
| Stunted | N (%) | 83 (15.1) | 91 (16.3) | 151 (12.5) | 207 (15.8) | 532 (14.7) | 58 (12.8) | 223,867 |
| Moderate stunted | N (%) | 78 (14.2) | 89 (15.9) | 136 (11.3) | 202 (15.4) | 505 (13.9) | 44 (9.7) | 169,475 |
| Severe stunted | N (%) | 5 (0.8) | 2 (0.4) | 15 (1.2) | 6 (0.4) | 28 (0.7) | 14 (3.1) | 54,393 |
| **Wasted and Stunted** | **N (%)** | 9 (1.6) | 5 (0.9) | 11 (0.9) | 20 (1.5) | 45 (1.2) | 3 (0.5) | 8,045 |
| **Reported small at birth<sup>‡</sup>** | **N (%)** | 46 (18.4) | 63 (22.9) | 87 (15.3) | 97 (16.1) | 294 (17.3) | 70 (14.8) | 258,847 |

<sup>‡</sup> mother's perception
<sup>‡</sup> All infants u6m were estimated using the following formula: $(\text{crude birth rate} \times \text{total population})/2$ ; $(0.0204 \times 171,467,000)/2 \approx 1,748,963$ infants u6m projected in 2023, Bangladesh. Source: World Population Prospects, 2024(Nations), available at https://population.un.org/wpp/ (accessed on 14 May 2025).

Of the 3,629 infants aged 0-5 months, 48.7% were female. The sex distribution was almost similar in all age groups. Infants were almost equally distributed across age groups, with the highest number in the 4-5-month age group (36.1%). Among the included infants, 12.6% (95% CI = 11.3; 14.1) were wasted, 14.7% (13.3; 16.1) were stunted, 15.9% (14.5; 17.3) were underweight, 1.24% (0.9; 1.7) were concurrently wasted and stunted (WaSt), and 17.3% (15.2; 19.6) were reported small at birth.

In the year 2022, 476 infants aged 0-5 months were included. Overall, sex distribution was almost equal (48.9% female). Among the included infants, 10.6% (7.9; 14.0) were wasted, 12.8% (9.5; 17.2) were stunted, 13.7% (10.6; 17.6) were underweight, 0.4% (0.1; 1.6) were concurrently wasted and stunted (WaSt), and 14.8% (11.7; 18.3) were reported small at birth.

**Table 2** also shows the caseload of malnutrition among all u6m old infants in Bangladesh. The country had an estimated total of 1,748,963 infants u6m. Among them, 185,390 were classified as wasted, 223,867 as stunted, 239,608 as underweight, 8,045 experienced both wasting and stunting, and 258,847 were small at birth.

**Figure 2** illustrates the mean values of WAZ, WLZ, and LAZ scores among infants of various age groups over two decades. WAZ had the lowest values among these three indices, while WLZ had the highest across all age groups. All three indices improved at the sixth month compared to the first month. However, all three indices declined during the 4 to 5-month period. WAZ and WLZ scores followed a similar trajectory throughout the 6-month period.

**Figure 2:**
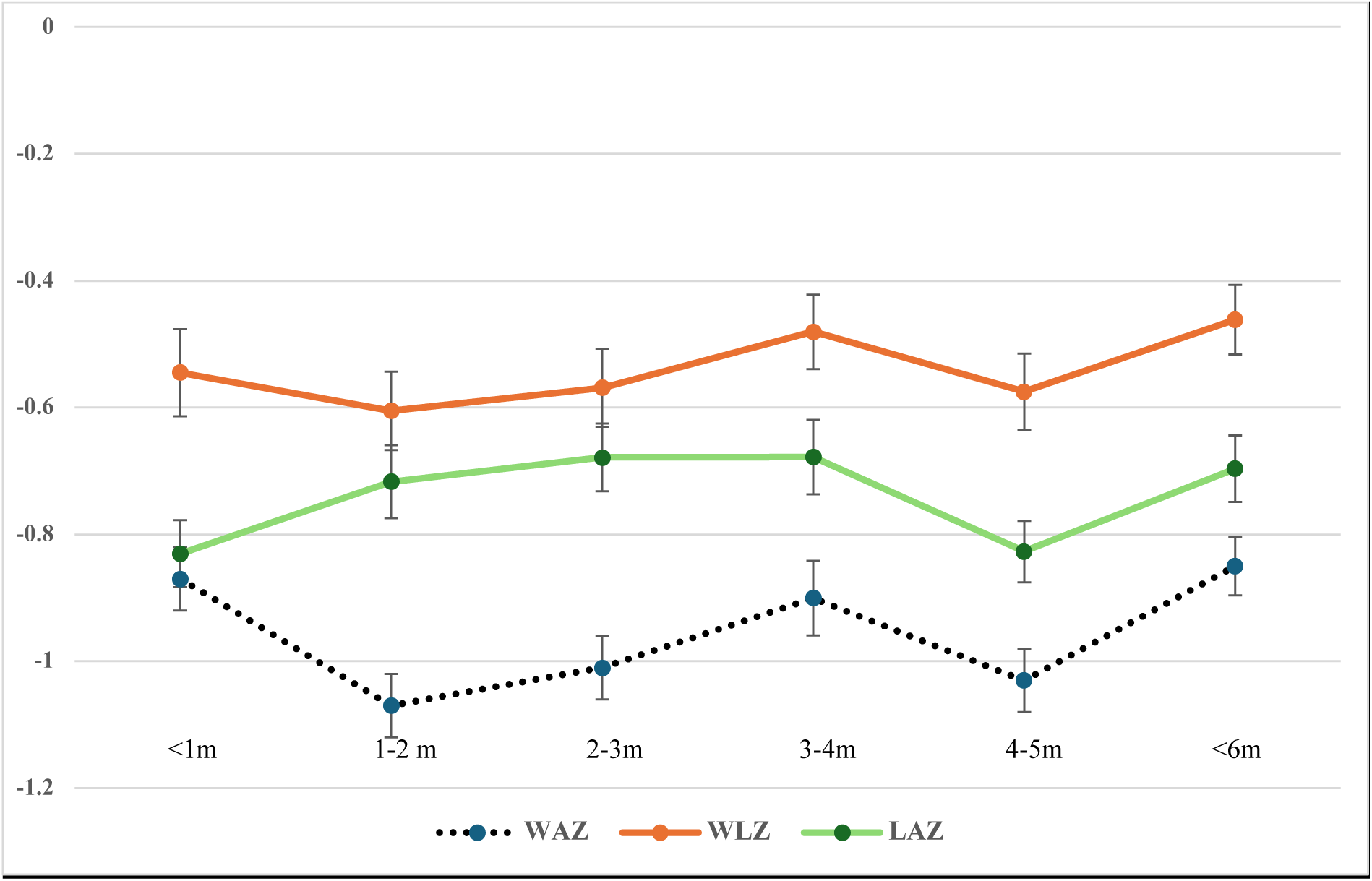
The trend of change in different anthropometric indices across infant age groups.

**Figure 3** provides an overview of the various forms of malnutrition among infants u6m and the overlaps between these conditions. It shows that underweight was the most common anthropometric deficit, followed by stunting and wasting. It also shows that all those concurrently wasted and stunted were also underweight.

**Figure 3:**
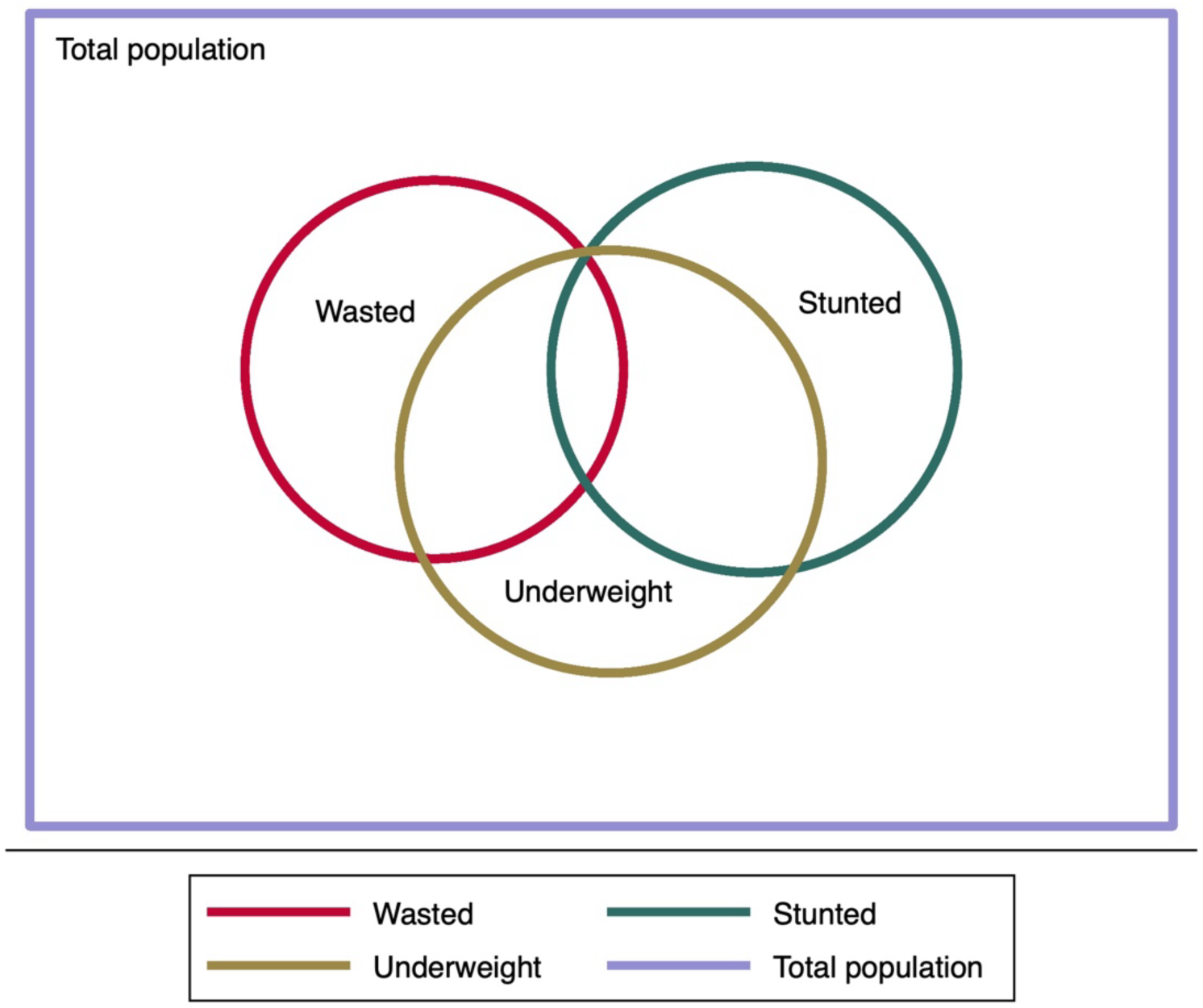
Venn diagram showing the prevalence and overlap of different anthropometric deficits.

**Figure 4** illustrates the prevalence of malnutrition across Bangladesh’s eight divisions over the last two decades. Overall, Mymensingh has the highest levels of malnutrition, whereas the Khulna division has the lowest.

**Figure 4:**
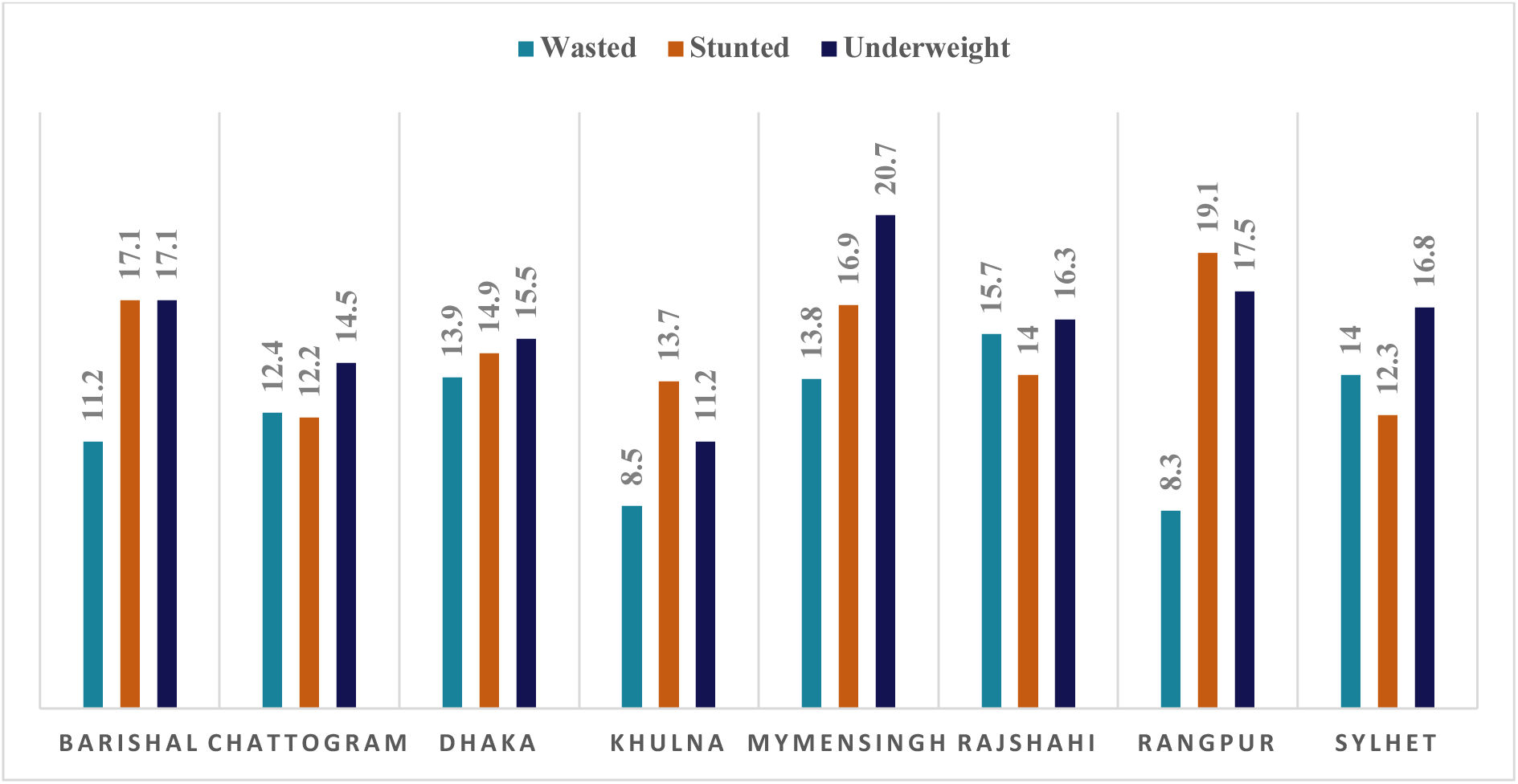
Wasting, stunting and underweight across regions/divisions of Bangladesh.

### 3.4 Anthropometric deficit over time

**Figure 5** illustrates the prevalence of different anthropometric deficits from 2004 to 2022.

**Figure 5:**
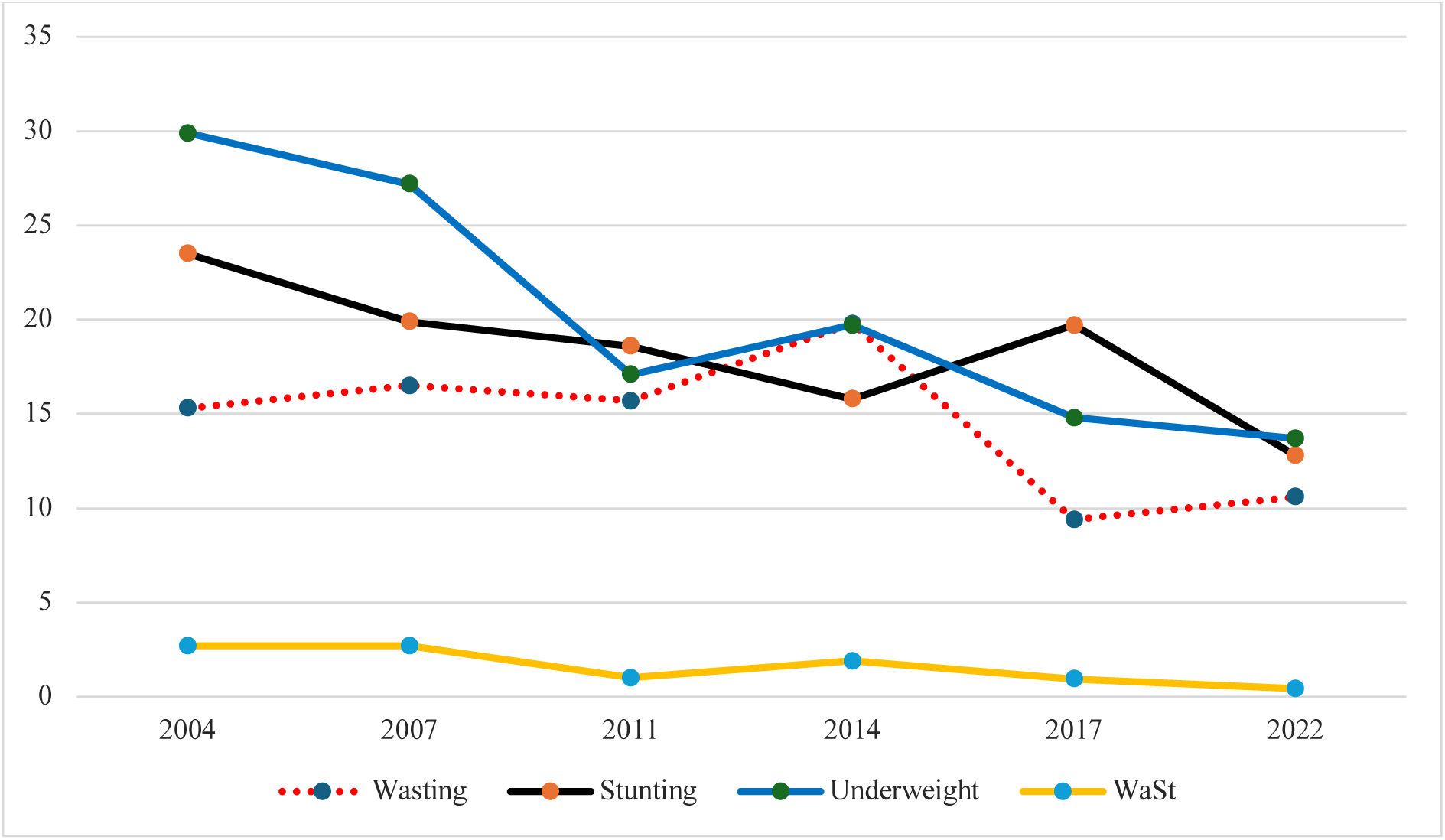
Prevalence (%) of wasted, stunted, underweight and WaSt infants from five datasets from 2004 to 2022.

Stunting declined from 2004 (23.7%) to 2014 (15.8%), and then increased to 19.7% in 2017. In the latest 2022 data, it decreased to 12.8%. The prevalence of underweight decreased steadily from 2004 to 2011. In 2004, the prevalence of underweight was 29.9%, compared to 17.1% in 2011. However, in 2014, it had increased to 19.7%; in 2017 and 2022, it again reduced to 14.8% and 13.7%, respectively. Wasting was static from 2004 to 2011, at around 15%. However, in 2014, it went up to 19.7%. Then, in 2017, it reduced to 9.4%, staying low at 10.6% in 2022. In 2014 and 2017, an inverse relationship was observed between wasting and stunting. In 2014, stunting prevalence fell to 15.8%. In contrast, wasting prevalence increased to 19.7%. On the other hand, in 2017, stunting prevalence increased to 19.7% and wasting prevalence decreased to 9.4%. Concurrent wasting and stunting was below 3% over the years. Overall, the rates of wasting, stunting, and underweight declined over the years, with increases in wasting and underweight in 2014 and in stunting in 2017.

### 3.5 Factors associated with wasted and underweight infants

**Table 2** shows the bivariate and multivariate analyses of different factors and their associations with wasting and underweight. In bivariate analysis, in terms of wasting, maternal BMI, maternal level of education, decision makers on maternal health, infant age categories, infant sex, place of delivery, exclusive breastfeeding, and availability of vaccine card were found to have a strong association. Mother’s age at delivery and maternal height showed some association with wasting. In terms of underweight, residence, type of toilet, cooking fuel, wealth index, mothers’s age at delivery, mother’s BMI, mother’s height, mother’s level of education, infant age categories, reported size at birth, birth order, number of ANC visits, the place of delivery, and presence of fever, cough and diarrhoea in the past two weeks were found to have an association.

In multivariable logistic regression, several factors were statistically associated with being wasted and underweight.

**Table 2:** Two anthropometric deficits, wasted (WLZ <-2) and underweight (WAZ <-2) and their associated factors.

| Risk Factor | Category | Wasted |  |  |  |  |  | Underweight |  |  |  |  |  |
| --- | --- | --- | --- | --- | --- | --- | --- | --- | --- | --- | --- | --- | --- |
|  |  | Unadjusted |  |  | Adjusted |  |  | Unadjusted |  |  | Adjusted |  |  |
|  |  | OR | 95% CI | P value | aOR | 95% CI | P value | OR | 95% CI | P value | aOR | 95% CI | P value |
| Household Characteristics |  |  |  |  |  |  |  |  |  |  |  |  |  |
| Residence (ref = urban) | Rural | 1.1 | 0.83 - 1.46 | 0.516 | 1.77 | 0.76 - 4.13 | 0.186 | 1.71 | 1.29 - 2.27 | <0.001 | 2.35 | 0.96 - 5.74 | 0.06 |
| Religion (ref = Muslims) | Hindus and others | 1.02 | 0.68 - 1.52 | 0.914 | 0.6 | 0.18 - 1.99 | 0.408 | 1.23 | 0.83 - 1.82 | 0.302 | 0.08 | 0.01 - 0.76 | 0.029 |
| Sex of the Household head (ref = Male) | Female | 1.31 | 0.89 - 1.93 | 0.175 | 2.74 | 1.02 - 7.39 | 0.046 | 0.78 | 0.54 - 1.12 | 0.179 | 0.85 | 0.28 - 2.60 | 0.781 |
| Type of toilet (ref = improved) | Un-improved | 1.01 | 0.76 - 1.33 | 0.947 | 0.74 | 0.36 - 1.51 | 0.405 | 1.48 | 1.18 - 1.85 | 0.001 | 1.39 | 0.66 - 2.92 | 0.387 |
| Cooking fuel (ref = Clean) | Unclean | 0.9 | 0.62 - 1.31 | 0.585 | 1.03 | 0.31 - 3.46 | 0.96 | 1.68 | 1.08 - 2.61 | 0.021 | 1.51 | 0.34 - 6.70 | 0.588 |
| Source of drinking water (ref = improved) | Unimproved | 1.31 | 0.67 - 2.50 | 0.413 | 2.02 | 0.46 - 8.96 | 0.352 | 1.22 | 0.68 - 2.20 | 0.499 | 4.24 | 1.04 - 17.3 | 0.044 |
| Wealth Index (ref = middle) | Poorest | 1.14 | 0.79 - 1.64 | 0.483 | 0.59 | 0.24 - 1.49 | 0.268 | 1.58 | 1.17 - 2.14 | 0.003 | 2.48 | 0.87 - 7.03 | 0.087 |
|  | Poorer | 1.1 | 0.78 - 1.56 | 0.578 | 0.62 | 0.22 - 1.74 | 0.364 | 0.97 | 0.71 - 1.32 | 0.841 | 0.57 | 0.19 - 1.71 | 0.315 |
|  | Richer | 0.93 | 0.64 - 1.34 | 0.691 | 0.66 | 0.26 - 1.70 | 0.389 | 0.6 | 0.43 - 0.85 | 0.004 | 1.27 | 0.32 - 5.02 | 0.732 |
|  | Richest | 1.27 | 0.89 - 1.80 | 0.193 | 2.66 | 0.94 - 7.54 | 0.066 | 0.79 | 0.55 - 1.13 | 0.199 | 3.53 | 0.76 - 16.4 | 0.107 |
| Maternal Characteristics |  |  |  |  |  |  |  |  |  |  |  |  |  |
| Mother's age (ref = <20 years) at delivery | 20-34 years | 1.14 | 0.86 - 1.52 | 0.357 | 2.09 | 0.72 - 6.06 | 0.175 | 1.24 | 0.99 - 1.56 | 0.065 | 1.92 | 0.50 - 7.40 | 0.342 |
|  | >=35 years | 1.56 | 0.93 - 2.62 | 0.089 | 1.71 | 0.62 - 4.68 | 0.296 | 1.64 | 1.02 - 2.66 | 0.043 | 2.38 | 0.84 - 6.75 | 0.103 |
| Maternal BMI (ref = normal) | Underweight | 1.52 | 1.15 - 2.01 | 0.003 | 1.77 | 0.82 - 3.83 | 0.148 | 2.19 | 1.71 - 2.79 | <0.001 | 2.05 | 0.93 - 4.51 | 0.074 |
|  | Overweight and obese | 0.82 | 0.58 - 1.16 | 0.257 | 0.68 | 0.27 - 1.72 | 0.414 | 0.63 | 0.44 - 0.91 | 0.015 | 0.17 | 0.05 - 0.62 | 0.007 |
| **Maternal height (ref = >145cm)** | **Height <145cm** | 1.17 | 0.84 - 1.63 | 0.353 | 0.89 | 0.29 - 2.71 | 0.84 | 2.34 | 1.79 - 3.05 | <0.001 | 5.03 | 2.11 - 12.01 | <0.001 |
| **Maternal level of education (ref = none)** | **Primary** | 0.72 | 0.49 - 1.04 | 0.079 | 0.55 | 0.24 - 1.26 | 0.159 | 0.51 | 0.37 - 0.70 | <0.001 | 0.69 | 0.30 - 1.59 | 0.391 |
|  | **Secondary** | 0.68 | 0.49 - 0.95 | 0.024 | 0.51 | 0.20 - 1.28 | 0.153 | 0.44 | 0.33 - 0.58 | <0.001 | 0.36 | 0.12 - 1.13 | 0.081 |
|  | **Higher** | 0.62 | 0.41 - 0.94 | 0.026 | 0.41 | 0.10 - 1.61 | 0.202 | 0.3 | 0.20 - 0.46 | <0.001 | 3.76 | 0.62 - 22.7 | 0.149 |
| **Mother's employment status (ref = Unemployed)** | **Employed** | 0.76 | 0.54 - 1.07 | 0.117 | 0.95 | 0.41 - 2.19 | 0.904 | 0.8 | 0.59 - 1.07 | 0.131 | 0.77 | 0.31 - 1.88 | 0.564 |
| **Total number of births (ref = less than 3)** | **3 or more** | 0.88 | 0.68 - 1.13 | 0.308 | 0.79 | 0.37 - 1.66 | 0.533 | 1.11 | 0.89 - 1.40 | 0.355 | 2.45 | 1.05 - 5.74 | 0.039 |
| **Infant Characteristics** |  |  |  |  |  |  |  |  |  |  |  |  |  |
| **Infant age category (ref <1 month)** | **1 to <2 months** | 0.89 | 0.61 - 1.31 | 0.56 | 0.48 | 0.14 - 1.63 | 0.239 | 1.64 | 1.13 - 2.38 | 0.009 | 1.24 | 0.35 - 4.39 | 0.74 |
|  | **2 to <4 months** | 0.82 | 0.59 - 1.14 | 0.234 | 0.93 | 0.37 - 2.30 | 0.873 | 1.38 | 0.97 - 1.98 | 0.076 | 0.89 | 0.26 - 3.09 | 0.864 |
|  | **4 to <6 months** | 0.65 | 0.46 - 0.92 | 0.014 | 0.54 | 0.18 - 1.59 | 0.265 | 1.46 | 1.02 - 2.07 | 0.038 | 0.7 | 0.18 - 2.67 | 0.602 |
| **Infant sex (ref = male)** | **Female** | 0.74 | 0.59 - 0.93 | 0.012 | 0.66 | 0.36 - 1.20 | 0.175 | 0.88 | 0.71 - 1.08 | 0.23 | 0.45 | 0.23 - 0.89 | 0.022 |
| **Reported size at birth (ref = average or large)** | **Small** | 1.32 | 0.85 - 2.04 | 0.217 | 1.05 | 0.46 - 2.38 | 0.899 | 4.18 | 2.79 - 6.28 | <0.001 | 6.13 | 2.71 - 13.8 | <0.001 |
| **Birth spacing (ref >36 months)** | **<=36 months** | 0.85 | 0.59 - 1.22 | 0.382 | 0.48 | 0.21 - 1.10 | 0.083 | 0.97 | 0.72 - 1.31 | 0.873 | 0.76 | 0.36 - 1.57 | 0.456 |
| **No. of ANC visits (ref = <4 visits)** | **>=4 visits** | 0.93 | 0.70 - 1.22 | 0.602 | 1.66 | 0.86 - 3.21 | 0.13 | 0.69 | 0.54 - 0.87 | 0.002 | 2.07 | 0.88 - 4.86 | 0.094 |
| **Place of delivery (birth) (ref = health facility)** | **Home** | 1.29 | 1.01 - 1.66 | 0.043 | 1.51 | 0.66 - 3.45 | 0.327 | 1.76 | 1.38 - 2.24 | <0.001 | 0.29 | 0.11 - 0.79 | 0.015 |
| **Postnatal check within 2 months (ref = no)** | **Yes** | 0.89 | 0.65 - 1.22 | 0.474 | 0.67 | 0.34 - 1.34 | 0.263 | 0.91 | 0.68 - 1.21 | 0.503 | 0.27 | 0.12 - 0.61 | 0.002 |
| **Initiation of breastfeeding (ref = less than 1 hour)** | **More than 1 hour** | 1.17 | 0.90 - 1.51 | 0.239 | 1.58 | 0.86 - 2.88 | 0.138 | 1.17 | 0.93 - 1.45 | 0.174 | 0.82 | 0.41 - 1.65 | 0.576 |
| **Exclusively breastfed (ref = no)** | **Yes** | 1.71 | 1.01 - 2.91 | 0.046 | 1.42 | 0.58 - 3.50 | 0.442 | 0.97 | 0.61 - 1.55 | 0.909 | 0.45 | 0.18 - 1.17 | 0.101 |
| **Infant given powdered, tinned or fresh milk on the previous day (ref = no)** | **Yes** | 1.23 | 0.64 - 2.36 | 0.538 | 0.75 | 0.22 - 2.58 | 0.646 | 1.46 | 0.80 - 2.68 | 0.214 | 1.88 | 0.67 - 5.27 | 0.231 |
| **Fever in past 2 weeks (ref = no)** | **Yes** | 1.07 | 0.85 - 1.35 | 0.582 | 1.59 | 0.72 - 3.48 | 0.247 | 1.49 | 1.19 - 1.86 | 0.001 | 1.98 | 0.1 - 3.97 | 0.052 |
| **Cough in past 2 weeks (ref = no)** | **Yes** | 1.11 | 0.88 - 1.40 | 0.361 | 1.46 | 0.69 - 3.07 | 0.315 | 1.29 | 1.03 - 1.61 | 0.028 | 0.68 | 0.32 - 1.45 | 0.318 |
| **Diarrhoea in past 2 weeks (ref = no)** | **Yes** | 1.11 | 0.57 - 2.16 | 0.765 | 0.73 | 0.18 - 2.94 | 0.657 | 1.66 | 0.97 - 2.83 | 0.062 | 0.52 | 0.13 - 2.06 | 0.352 |
| **Vaccine card present (ref = no)** | **Yes** | 0.77 | 0.60 - 0.98 | 0.037 | 0.8 | 0.38 - 1.69 | 0.561 | 0.97 | 0.77 - 1.23 | 0.822 | 1.37 | 0.61 - 3.05 | 0.318 |

### Household Characteristics

Children from non-Muslim households were 92% less likely to be underweight compared to those from Muslim households (p = 0.029). Children from households with an unimproved source of drinking water were 4.24 times more likely to be underweight than the improved drinking water source counterpart (p = 0.044). Additionally, infants from female-headed families were 2.74 times more likely to be wasted than male-headed families (p = 0.046). Some evidence indicates that infants of rural households were 2.35 times more likely to be underweight than those from urban households (p = 0.06).

### Maternal Characteristics

Infants born to mothers with a BMI indicating overweight and obese were 83% less likely to be underweight, compared to those born to mothers with a normal weight (p = 0.007). Additionally, infants of underweight mothers had 2.05 times higher odds of being underweight compared to those of mothers with a normal weight (p = 0.074); however, the evidence for this finding is not strong. A strong association was observed between maternal height and infant underweight status. Infants whose mothers had a height of less than 145 cm had a 5.03 times higher chance of being underweight than those with taller mothers (p < 0.001). Additionally, mothers who had three or more births had 2.45 times higher odds of having an underweight infant, though the evidence was weak (p = 0.039).

### Infant Characteristics

Female infants had a 55% lower chance of being underweight than their male counterparts (p = 0.022). Infants’ reported size at birth was a significant factor in the likelihood of underweight. Infants classified as small at birth were 6.13 times more likely to be underweight (p < 0.001) than infants of average or larger size at birth (p < 0.001). The place of delivery also showed a significant association with child health outcomes. Home-based deliveries were linked to a 71% lower risk of being underweight (p = 0.015) than hospital-based deliveries. Another factor found to be associated with underweight was receiving PNC services. Infants whose mothers received a PNC check-up within 2 months of delivery had a 73% lower chance of being underweight than infants whose mothers did not receive any PNC in that timeframe (p = 0.002). Infants who had a fever within the past 2 weeks were found to be associated with underweight, showing 1.98 times higher odds of being underweight compared to those without fever (p = 0.052). Some evidence indicates that infants born with a 36-month birth spacing had a 52% lower chance of wasting (p = 0.083). In the adjusted analysis, infants age category, exclusively breastfed children, cough, diarrhoea, or having a vaccine card showed no significant association with wasting or being underweight.

## 4. DISCUSSION

In Bangladesh, DHS survey data indicate that, however assessed, infant u6m was common. Over the last two decades, 12.6% of infants 0-5 months were wasted, 14.7% were stunted, and 15.9% were underweight. It remains a common problem based on the latest 2022 data, with 10.6% wasted, 12.8% stunted, and 13.8% underweight. Several observations support calls from other research to focus on underweight as the preferred measure of malnutrition in this age group. Firstly, there is data quality: length-based measures are overall poorer, with LAZ and WLZ having more flagged data. Underweight was also more strongly and more consistently associated with a number of biologically plausible risk factors for malnutrition: rural residence, religion, high-risk drinking water source, low maternal BMI, low maternal height, total number of births, infant sex, reported size at birth, delivery place, PNC, and fever in the past 2 weeks. For wasting, only sex of the household head was found to have a significant association.

In this study, infant u6m malnutrition is a common problem in Bangladesh, supporting the need for treatment and prevention programming, consistent with what the latest WHO Malnutrition guidelines recommend. This status of malnutrition among infants u6m is, however, better than the overall world’s prevalence (Kerac et al., 2025). Across different anthropometric measurements, LAZ had the highest number of flagged data (259, 6.4%), followed by WLZ (6.0%) and WAZ (0.8%). Other studies had similar findings (Grijalva-Eternod et al., 2017; Mahmud et al., 2024). Studies have indicated that length is particularly prone to measurement errors due to the natural/infantile position of the legs, which must be straightened during measurement (Mwangome & Berkley, 2014). This makes the measurement more complicated and time-consuming for the health worker and may be less acceptable for the mother, as it requires moving the infant into this position (Lelijveld & Kerac, 2017). Another critical finding was that those who were concurrently wasted and stunted were also underweight. A similar finding was observed in other studies as well (Mahmud et al., 2024; Myatt et al., 2018).

In terms of wasting, stunting and underweight trends, improvements over the last 20 years were observed. The highest progress was observed for underweight, while the prevalence of wasting has declined modestly. This progress may have been possible because of the government’s multisectoral interventions focusing on both nutrition-specific and nutrition-sensitive interventions (BNNC, August 2017; Suri, 2019). Regional disparities were observed across Bangladesh, with Mymensingh as the most affected division, showing the highest level of underweight. Addressing malnutrition among this age group might require prioritisation and a targeted approach to the most affected regions.

Several factors were found to be associated with wasting and underweight. This indicates a need for packages of care which explore and address multiple risk factors rather than one alone (Grey et al., 2021). Reported small size at birth was, for example, found to be positively associated with the underweight status of the infants. Small size at birth can result from conditions like intrauterine growth retardation (IUGR) or preterm birth (De Bernabé et al., 2004). This observed association is consistent with other studies (Rahman & Chowdhury, 2007; Rahman et al., 2016; Rayhan & Khan, 2006). This supports WHO 2023 guidelines to treat these infants as “at-risk of poor growth and development” since infants born small tend to remain undernourished throughout their whole infancy (Arifeen et al., 2000) and remain vulnerable to various diseases and complications (Akdemir, 2010; Anand et al., 2003; Romero et al., 2012). It highlights the importance of pre-pregnancy and antenatal nutrition of mothers since child malnutrition onset from the foetal period (Fall, 2013; Nemoto & Kakinuma, 2020).

Female infants were found to be less prone to develop underweight than their male counterparts. This finding is well supported in both regional and international studies (Khara et al., 2018; Mahmud et al., 2020; Susan et al., 2020). This can be explained by the early-life vulnerability of male children; in contrast, female children often show better physiological and immunological resilience (Naeye et al., 1971; Wells, 2000). This may contribute to a better nutritional status in female infants. Although in some societies, such as Bangladesh, females are often neglected within the family in terms of food allocation and care-seeking behaviours, this may be neutralised during infancy, when biological differences mostly dominate (Chen et al., 1981).

Strong evidence was found for an association between the place of delivery and infant underweight. Infants delivered at home had higher odds of being underweight compared to infants delivered at a hospital, but adjusted analysis shows lower odds. This contradicts other evidence indicating that institutional delivery is better for health outcomes (Olusanya & Renner, 2012). A likely explanation is selection bias. In Bangladesh, many people view pregnancy as a natural process and prefer not to go to a hospital unless complications arise. They prefer home delivery over hospital due to traditional views, religious considerations, limited access of women to decision-making in the family, unaffordability, lack of transportation to reach the nearest health facility and to avoid caesarean section (Sarker et al., 2016). Other unmeasured confounders, such as the quality of facility-based care or home birth-related cultural practices, might also affect this finding. Our observation certainly does not imply that home delivery protects against infant malnutrition; instead, it highlights the need to do further research on who delivers at home vs in hospital and on the quality of facility-based healthcare services.

In our analysis, maternal postnatal check-ups within two months of delivery were associated with less underweight. This is biologically plausible since mothers thus presenting are also likely to be more proactive, engaged and empowered (Hamel et al., 2015; Pokhrel et al., 2016; Sigdel et al., 2020). During PNC services, mothers receive vital information on exclusive breastfeeding, vaccination, hygiene, and proper infant care. This knowledge also contributes to reduced infant malnutrition.

Maternal BMI was associated with underweight. Infants of overweight or obese mothers had an 83% lower chance of being underweight compared to the normal-weight mothers. This concurs with other surveys (Casapia et al., 2007; Dekker et al., 2010; Felisbino-Mendes et al., 2014; Hasan et al., 2016; Rayhan & Khan, 2006). There is, however, also a risk that these children may develop overweight, obesity and non-communicable diseases in early childhood or later in life (Agras et al., 2004; Salsberry & Reagan, 2005).

Short maternal stature (under 145 cm) was associated with infant u6m underweight. This is also biologically plausible since maternal height is an indicator of intergenerational risk as well as a mother’s own childhood nutrition and health. A secondary analysis of BDHS data showed that maternal short stature was associated with 1.3- and 1.4-times higher risks of wasting and severe wasting, respectively. This study also found that a one-centimetre increase in maternal height was associated with a 1.4 % decrease in wasting (Khatun et al., 2019). Shorter maternal stature, an indicator of chronic undernutrition, leading to suboptimal nutrient stores, impaired fetal growth and underweight in infancy (Khanam et al., 2019; Kozuki et al., 2015; Muhihi et al., 2016; Rush, 2000) (Hart, 1993; Van Roosmalen & Brand, 1992). Also, short maternal stature is linked to poverty, limited access to healthcare and poor living conditions, which can contribute to underweight infants. In our analysis, we indeed found a significant association with wealth quintiles and both wasting and underweight infants.

In this analysis, infants of non-Muslim families were found to have lower odds of developing underweight compared to their Muslim counterparts. This does not align with some other studies (Banerjee & P, 2023). It shows that infant undernutrition is complex and associated with social structure, lifestyle, dietary habits, etc.

We also revealed a link between female-headed households and infant wasting. This mirrors others’ findings (Ashagidigbi et al., 2022). In Bangladesh, most families are male-headed, and female-headed households often consist of widows or single parents who lack a male provider to assist with generating income. Often, women’s wages are lower than those of their male counterparts, which may further affect infant nutritional status (Rahman & Al-Hasan, 2019). Being the head of the household might also lead to increased need to work outside, which affects their ability to spend time with, care for and breastfeed their babies (Hasan et al., 2020).

Consistent with other studies, our data showed that unimproved drinking water sources were associated with increased infant underweight (Braghetta, 2006; Soboksa et al., 2021). Mechanisms might include increased risk of enteric infections, leading to malnutrition. Fever in the last two weeks was also associated with underweight and a marker of infections, which directly and indirectly affect infant growth (Weisz et al., 2011). The practical implication of the finding is that it is important to support infant health and not just nutritional intake.

### 4.1 Strengths and limitations

Since we rely on observational, cross-sectional data, we acknowledge several limitations to our work. The key risks are unknown confounding and bias; these should be considered when interpreting our results. Even when biologically plausible, associations between risk factors and WLZ and WAZ should be interpreted cautiously, as hypothesis-generating. Secondly, there may be misclassification of exposure due to recall bias when interviewing the mother. This may be less of a problem for infant u6m-related information since the time lag is less than six months. It may, however, be problematic when a mother replies with socially desirable answers; this might explain why breastfeeding did not emerge as a protective factor in our analysis, as might have been expected. Mothers may have answered positively while not actually breastfeeding. Measurement bias is another challenge. Although the interviewers were trained similarly and given the same information for interviews and measurements, there may still be variable responses across interviewers and inaccurate measurements. The choice of DHS dataset may also introduce selection bias. The children’s dataset was used, which included infants whose mothers were home and eligible for an interview. The study did not include infants and mothers who were not at home during the household survey. This could mean that the mother and/or infant were hospitalised during the survey and have poorer health and nutritional outcomes. Another reason for a mother not to be present could be that the infant was an orphan or that the mother was working. These children may have a different nutritional status than the children with an interviewed mother. Not including these infants in our analysis may mean our prevalence estimates are lower than the true prevalence, especially for severe wasting, which often requires inpatient treatment (WHO, 2024). In this analysis, we combined data from 2004 to 2022 and assessed various factors associated with infant malnutrition. It would have been ideal to evaluate factors for each dataset individually. However, due to the insufficient sample size, we were unable to do so. Nonetheless, we do not think it will change the actual message we wanted to convey.

Despite the limitations, this study has some strengths. To our knowledge, this is the first study that provides an estimate of the total caseload of different forms of malnutrition among u6m old infants in Bangladesh. This will be useful to local policymakers and programme managers. The basic analytical framework used in this study can be easily and rapidly replicated for other countries seeking to better understand the local epidemiology of infant u6m malnutrition. The generalizability of the approach to similar settings is thus high. Another strength of DHS data is that all the measurements were taken from a representative population sample through a household survey with a high response rate and a two-stage cluster sampling strategy. No intervention or new measurement technique has been introduced in this study. Therefore, the results reflect the current ‘real life’ situation.

### 4.2 Future study

For future research, it would be important to examine differences in the associations between moderate and severe wasting or underweight with different risk factors. Ideally, the association between WLZ and WAZ should be assessed with mortality in a prospective study in a community setting. So far, this has only been done in a clinical setting (Mahmud et al., 2024; Mwangome et al., 2017). A prospective study can also help us better understand the risk factors associated with malnutrition in infants. After checking their anthropometric indicators, follow-up of these infants is needed to understand the mortality and nutritional risk for wasted infants who would not be included in the nutritional intervention when WAZ is used instead of WLZ. The reason why the prevalence of wasted and underweight infants increased between 2011 and 2014 needs to be evaluated.

## 5 CONCLUSIONS

Over the past two decades, Bangladesh has markedly reduced various forms of undernutrition among infants u6m. However, the prevalence is still high. Our analysis highlights the importance of focusing not just on nutritional intake but also on wider infant, maternal and social factors which may underlie undernutrition. To effectively address this issue, packages of care are needed as well as prospective studies to identify which nutritional interventions will most effectively treat and prevent malnutrition in infants.

## Supporting information

S1 Table: Distribution of the sample population in eight districts of Bangladesh

## Data Availability

All data produced are available online at DHS website (https://dhsprogram.com)

## Acknowledgments

We would like to express our gratitude to the Demographic Health Survey (DHS) Program for providing data access used in this research. We are also thankful to Dr Md. Munirul Islam for the initial conceptualisation of the research.

## Data Availability Statement

Publicly available data were used, which was accessible from the DHS website (https://dhsprogram.com) upon request.

## Authors’ contributions

MK, IM, RA, ABS, KTR, and RH conceived the study. IM conducted data curation and formal analysis. IM, RA, MK, and ABS contributed to the methodology. IM and RA wrote the original draft. IM worked on data visualisation. MK supervised the whole study. All authors contributed to the manuscript revision. All authors read and approved the final draft.

## Supporting Information

Additional supporting information can be found online in the Supporting Information section.

**Table S1: Distribution of the sample population in eight districts of Bangladesh**

