## Supplementary material for "Prevalence, trends, and determinants of malnutrition among under six-month-old infants in Bangladesh: analysis of DHS data (2004 – 2022)": S1 Table: Distribution of the sample population in eight districts of Bangladesh

| **Division** | **Year 2004 - 2022** | |
|---|---|---|
| | **Frequency** | **Percentage** |
| Barisal | 224 | 6.2 |
| Chattogram | 837 | 23.1 |
| Dhaka | 1,038 | 28.6 |
| Khulna | 367 | 10.1 |
| Mymensingh | 479 | 13.2 |
| Rajshahi | 358 | 9.9 |
| Rangpur | 234 | 6.4 |
| Sylhet | 92 | 2.5 |
| Total | 3,629 | 100 |
